# Design of optical cavity for air sanification through ultraviolet germicidal irradiation

**DOI:** 10.1101/2021.03.04.21252517

**Authors:** Matteo Lombini, Emiliano Diolaiti, Adriano De Rosa, Luigi Lessio, Giovanni Pareschi, Andrea Bianco, Fausto Cortecchia, Mauro Fiorini, Giulia Fiorini, Giuseppe Malaguti, Alessio Zanutta

## Abstract

The transmission of airborne pathogens represents a major issue to worldwide public health. Ultraviolet light irradiation can contribute to the sanification of air to reduce the pathogen transmission. This study concerns the design of a compact filter for airborne pathogen inactivation by means of UV-C LED sources, whose effective irradiance is enhanced thanks to high reflective surfaces. Ray-tracing and computational fluid dynamic simulations are both used to model the device and to maximize the performance inside the filter volume. Simulations foresee the inhibition of SARS-Cov 2 also in the case of high air fluxes. This study demonstrates that current available LED technology is effective for air sanification purposes.

## 1. Introduction

The coronavirus SARS-CoV-2 [1] pandemic has spread worldwide since beginning of 2020. There is evidence that aerosols, (i.e., small airborne particles and micro-droplets in the size range up to 5 *µ*m diameter), are a significant transmission pathway [2–4] and some case studies indicate that SARS-CoV-2 has viable survival rates in the air and it remains airborne for several hours [5, 6]. More generally, airborne transmitted microorganisms represent one of the major challenges to worldwide public health [7], as for example the seasonal or pandemic influenza [8–10]. Ultraviolet Germicidal Irradiation (UVGI) can efficiently inactivate airborne pathogens as well as drug-sensitive and multi-drug-resistant bacteria [11] and various strains of viruses [12]. The pathogens inactivation by means of Ultraviolet (UV) light started from the work by Downes and Blunt [13, 14]. Different reviews on the history of UVGI [15, 16] report on the available UV sources; moreover, the mechanisms of pathogens inactivation by UV light have been deeply investigated. UV-C photons, namely between 220 and 280 nm in wavelength, are absorbed by DNA/RNA and three major damages can occur: pyrimidine dimerization, which is the dominant damage, pyrimidine (6-4) pyrimidone photoproducts formation and protein-DNA cross-linking [17]. In pyrimidine dimers, two adjacent thymines in DNA/RNA produce a thymine-thymine dimer, disrupting the ability of the microorganism to replicate and thus to pose infection [18, 19]: they are called inactivated microorganisms. The inactivation rate depends on the irradiation wavelength [20]. Looking at the absorption spectrum of nucleic acids and proteins, the formers show a peak around 265 nm, whereas the latter show an absorption increase at wavelengths shorter than 245 nm and surpasses DNA absorption at 225 nm [21,22]. Moreover, depending on the protein structure, its UV absorption can be observed also in the UV-B and UV-A. Due to these absorption features by proteins, other types of damage can occur, such as the disruption of the cell membrane and consequently cell death or the denaturation of the capsid proteins in viruses with the infection prevention. Although it is known that this kind of UVC radiation has an inactivating effect on all microorganisms and viruses, all pathogens require different UVC irradiation doses for successful inactivation [15, 23]. Anyway the inactivation rate seems to be independent of the pathogen concentration [24]. High UV fluxes are important for different reasons: given the size of the protein UV absorption, the damage to the cell membrane occurs when the cell is exposed to a high UV fluence [18]; studies showed that microorganisms inactivation happened when of damaged sites on the DNA of microorganisms, proportional to the UV flux intensity, reaches ≈ 100 [25]; some microorganisms can repair the damaged sites through either dark or photoreactivation repair mechanism [26] if they are subjected to low fluxes.

Regarding the airborne pathogens, from the quite far in time works describing the inactivation through UVC irradiation [27, 28], many efforts have been done toward the so called bio aerosol control [29] both regarding the techniques [15,30] and the measurements of reference inactivation dosages [31, 32]. Besides the species peculiarities, the inactivation doses for aerosol applications depend also on relative humidity (RH), since there are evidences of microorganisms susceptibility increase with decreasing RH [33, 34], and on the effective filtration of dust [35], which absorbs light and shields pathogens.

The SARS-Cov-2 pandemic boosted the research and applications regarding the pathogens inactivation by means of several light based technologies [36]. The most used sources for UVGI applications are the mercury vapor lamps, for surfaces, water and air sanification. These sources emit UV radiation when an electrical current in the gas excites mercury vapor atoms trapped in the quartz envelope (mainly cylindrical). Based on the pressure of the mercury vapor in the envelope, they can be divided into low pressure (LP) and medium pressure (MP). The principal emission wavelength of LP lamps is 253.7 nm, while MP lamps have a broad emission spectrum (185–600 nm). During the Minamata convention on mercury in 2013 [37], organized by an United Nations Environment Program (UNEP), it was decided to forbid by 2020 mercury-containing merchandises for the well-being of human and environmental health. Nevertheless, mercury UVC lamps are still used in UVGI systems thanks to their sufficiently high emission power with reasonable costs. It is apparent that the use of alternative and more environmental friendly UVC sources is necessary. UVC Light Emitting Diodes (LED) are surely the best candidates [38]. A LED is a solid-state semiconductor device that emits light when is connected to direct current. Such light source shows important advantages, such as a low driven voltage, a very fast response time, a straightforward intensity control and it does not contain dangerous heavy metals (in particular mercury) [39]. UVC LEDs do contain small amounts of elements such as the metals gallium and magnesium and the metalloids silicon and boron (although boron is not predominantly used), which are bound within a stable crystal structure and cannot leach into the environment. The emitting region of an LED is about 1 *mm*^2^, thus for most applications they can be considered point-like sources. The emission spectrum resembles a Gaussian profile [40], with a usual Full-Width at Half Maximum (FWHM) of about 10–15 nm. In 2014, the first deep ultraviolet LEDs were brought to the market, after a development by Isamu Akasaki and Hiroshi Amano (awarded with Nobel Prize for Physics in December 2014), and are now available in multiple wavelengths within the germicidal range (255, 265, 275 nm, among others) [41]. The peak wavelength names conventionally an UV-LED. Current devices deliver UVC emission power values in the 50 to 100 *mW* range, with lifetime to 70% of initial output (L70) of around 10,000 hours. Different studies have already quantified the sanification effectiveness operated by LED sources for surfaces [20, 42], water [43] and air [32, 44].

This paper deals with the design of a compact filter for the sanification of air through UVC light. Despite the currently available LED sources emission power is of the order of tens to few hundreds of milliWatts, to be compared to the mercury vapor lamps emission of several watts, highly reflective surfaces combined to some shrewdness of the optical cavity design permit to have a compact and effective device. The same optical concept, where reflecting surfaces and a proper design of the filter shape are combined to maximize the internal energy density, can be applied to bigger fluxes of air, as for industrial air ducts, where anyway the required emission power can be reach, at the moment, only by mercury vapor lamps [45]. The concept of power density enhancement inside a volume due to a high reflectivity of the internal surfaces is widely used in many technological applications, such as integrating spheres [46] or Fabry-Perot interferometers [47]. Regarding the UVGI, Jensen in 1964 [48] describes the increase of sanification efficiency by an UVC source inserted in a reflective tube and similar experiments can be found in more recent works [44, 49, 50]. Since no secondary effects are produced after light absorption by pathogens (or air), the total UVC light total dose can be administered ‘in pieces’ after any of the numerous internal reflections, accordingly to the Bunsen and Roscoe law [51].

A possible application of the proposed UVC light compact filter can be the sanification of the exhaled air of infectious hospitalized patients treated with assisted ventilation, by means of continuous positive airway pressure (CPAP) or non-invasive ventilation (NIV) helmets where the size and weight constraints are important. Since the exhaled aerosol to the room can cause risk of viral transmission to healthcare workers [52], disposable Heat and Moisture Exchangers (HME) filters are placed before the Positive End-Expiratory Pressure (PEEP) valves. The efficiency of standard devices as the HME filters has been investigated [53–55]. They must be disposed after the use, about 24 hours time-scale, while the presented UVC light filter would be an effective, long lasting and self cleaning replacement. Standard working air flows for assisted ventilation devices are up to 100 l/min, which is the value to which filter efficacy simulations are referred to. The day that UVC LEDs will be more powerful and more efficient, even more compact (portable) sanification devices or bigger air flows could be treated.

## 2. Filter design

Regarding the UVC sanification of surfaces, the usual unit of measure is the *irradiance*, which is defined as total radiant power from all directions incident on an infinitesimal element of surface area *δ*S containing the point under consideration, divided by *δ*S, with typical SI units of *mW* / *cm*^2^ [56]. Irradiance is generally used, in the framework of air sanification, when the arrival directions of the light is mono directional [35, 57] or, in case of different sources, where the sensors for the light intensity measurement can be oriented toward the sources and an average facial irradiance is calculated [34, 58]. When light is coming from different directions, a more appropriate unit of measure is the *fluence rate*, defined as the radiant power passing from all directions through an infinitesimally small sphere of cross-sectional area *δ*A, divided by *δ*A, with typical units of *mW* / *cm*^2^. This is the case when reflecting and scattering surfaces homogenize the light travel directions [44, 49]. *Fluence* is the total radiant energy from all directions passing through an infinitesimally small sphere of cross-sectional area *δ*A, divided by *δ*A, with typical units of *mJ* / *cm*^2^. Fluence is also sometimes called UV dose, since it is the product of the fluence rate times the exposure time in seconds. For purposes of calculation volumes can not be considered infinitesimal, but as long as the areas are small, irradiation and fluence rate are used in their finite differences, substituting *δ*A with ΔA, which is a good approximation. Anyway, in this case, some rays do not pass through the sphere center. Some considerations regarding the fluence rate estimation for this case are discussed in Section 3.3.

### 2.1. Geometrical considerations

The filter for air sanification presented in this paper is a device composed by: a cavity, which has the function of enhance the sources power density and confines as much as possible the light inside its volume; two flanges, which can be connected to the inlet and outlet air pipes (Figure 1). The light emitted from the LED sources, placed at the edges of the internal volume of the filter, is bounded as much as possible inside the filter itself. The filter has a cylindrical shape even though different geometries could be used to optimize the performance. Rounded cavity profiles could lead to a filter efficiency optimization, both regarding internal fluence rate increase and the light leak reduction through the holes. The manufacturing process is more difficult and some investigations should be carried out to measure the highly reflective coating behavior when the surfaces are rounded. Moreover, the LED array plate has to be customized to follow the surface curvature. Further investigations will be accomplished to understand if these solutions are worth, considering performance and mass production requirements.

**Fig. 1.**
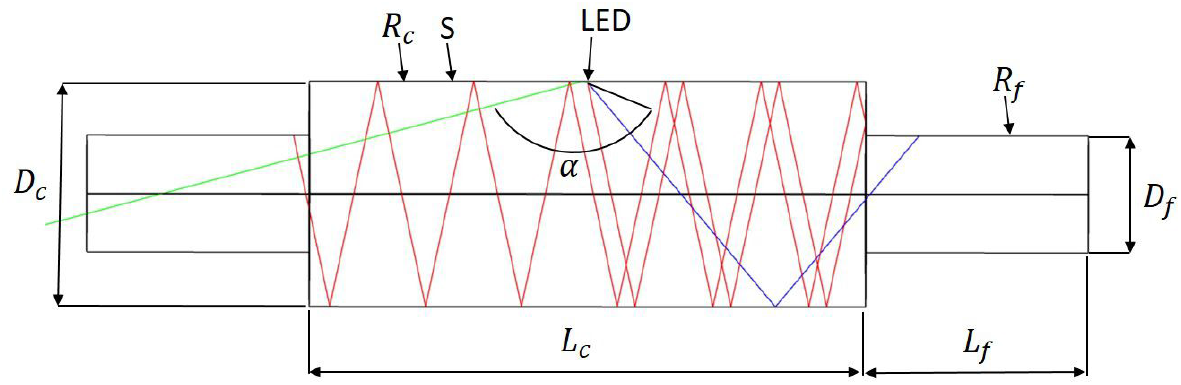
Sketch of the longitudinal section of the filter and indication of the parameters that are varied in the optical simulation trade-off, described in Section 3. Symbols legend: *R*_*c*_ cavity reflectivity; *D*_*c*_ cavity diameter; *L*_*c*_ cavity length; *L* _*f*_ flange; length; *R* _*f*_ flange reflectivity; *Sc* Scattering distribution; *α* LED emission cone angle

### 2.2. Light sources

The emission angle pattern of a given LED is provided in the manufacturer specifications documents and is usually defined in polar coordinates. The most common LED emission patterns vary with the polar angle *α* but are uniform in the azimuthal direction. Among the possible commercial LEDs, the optical simulations described in Section 3 consider the 6060 SMD model from Laser Components^®^ [59], which has 6×6 mm chip size, 100 mW radiant flux, 272 nm central wavelength and an angle *α* of 150°. The emission profile shown in the normalized red curve of polar plot in Figure 2 has been used for the optical simulations. LED emission angle can be reduced by means of a micro-lens with cavity mounted over the chip [60]. The micro-lenses must be glued to each LED and disposed of when the sources must be replaced. Some light loss is due to internal reflections and quartz absorption but the emission angle is reduced down to *α* ~ 30°. The blue curve in the normalized polar plot of Figure 2 shows a simulated emission angle pattern for a LED with micro-lens. There is a big impact on the power density distribution inside the cavity, as displayed in Figure 2 right and quantified in Section 5.2.

**Fig. 2.**
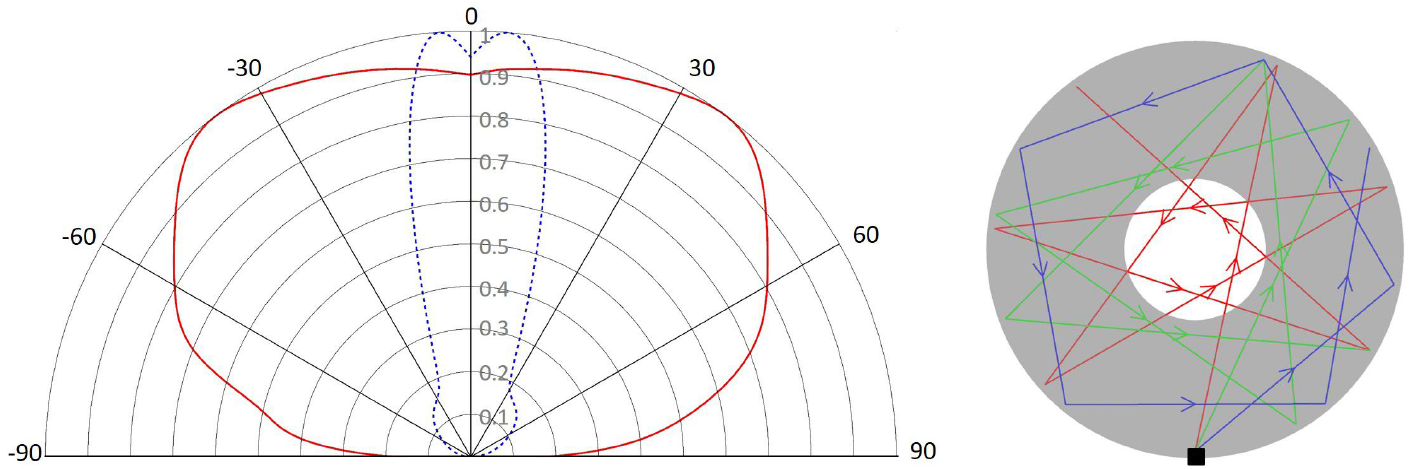
Left: normalized LED radiation profile in polar coordinates for the two different emission cone angles used in the simulations. The half emission angle *α* resents the angle at which the intensity of radiation becomes half of the maximum intensity; right: paths of different rays emitted by a point source inside a reflecting circle. The ray emission angle is maintained after any specular reflection until all the ray energy is totally absorbed or, in the case of the optical simulations, until the energy drops below a defined threshold.

A simplified geometrical explanation of the rays distribution inside the filter as a function of the emission angle is displayed in Figure 2 right, which is a plane perpendicular to the cavity axis. A point source representing the LED emits some rays, lying on the plane, which are reflected by the cavity inner surface. The inner circle represents the projection of the volume where almost all the air flow is confined, as indicated in Section 4, for the considered velocities. Due to Snell’s law, in case of specular reflection the angle of an emitted ray respect to the surface normal (direction from the source to the center), will be maintained for every reflection. Thus, the emitted rays that skip the inner circle at the launching, will continue to skip it after any reflection, and consequently they will not contribute to the power density in that region. Sources with a large emission angle as the reference LED emit more rays outside the inner circle while narrower emission angle sources make a bigger rays percentage to cross the circle. The actual rays distribution is also dependant on the cavity diameter. Scattering reduces or even cancels the emission angle pattern, mixing the rays directions and homogenizing the power density distribution, as shown in Figure 6 h.

#### 2.2.1. Heat dissipation

The power conversion for the UVC LEDs is still very low compared to devices optimized for other wavelengths. Another relatively new LED technology is the blue LEDs developed recently, also for TV screen; these devices have a power conversion efficiency (optical-per-electrical) that reach 80%, while this value for UVC LEDs is still below 10% [61]. So most of the electrical power is converted to heat rather than photons and some means are needed to dissipate it, to ensure the device will not be destroyed; a LED also will have a shorter life if it works constantly at higher temperature than expected. Heat can be removed primarly by thermal conduction along the Printed Circuit Board (PCB), the mechanical support of the LEDs and the other circuitry. Metal Core PCBs are boards which incorporate a metal layer as heat spreader inside the thickness of the board, usually consisting of a core of aluminium alloy. This is not enough for high current devices, so a heat sink must be used on the other side of the PCB hosting the LEDs, with a thermal adhesive to assure the maximum heat transfer between the board and the heat sink. In cases where the produced heat is higher, or when the natural convection between the fins of the heat sink is not enough to spread the heat, a forced air flow must be imposed with fan(s) directly attached to the heat sink.

### 2.3. Reflectivity and scattering

As already explained, the filter concept relies on the fluence rate magnification inside the cavity due to the multiple reflection of the rays from very reflective surfaces. The magnification or enhancing factor for a closed cavity can be calculated from the material reflectivity *R*. Given a ray with power *p*_0_ when launched from the source, after every reflection the power will be reduced by a factor *R* and after *n* reflections will be *p*_0_ · *R*^*n*^. In the hypothesis that all the light remains confined inside the optical cavity, the ray power contribution to the overall internal power density will be:

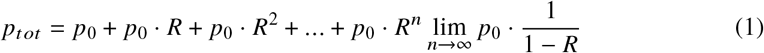

In reality some light leaks the cavity or is absorbed by the flanges or hit the (almost) non reflective LED package, thus the enhancing factor is reduced respect to eq. 1, but still the internal surfaces reflectivity is a fundamental parameter for the filter efficiency, as demonstrated in Figure 5.

The simulations reference case (Table 1) is considered to have a perfectly specular internal reflection. This ideal case is anyway plausible, or at least not very different, when the rays diffusing cone angle is narrow or the scattered rays fraction is small. The scattering of the internal cavity surfaces causes the fluence rate distribution inside the volume to be smoothed respect to a pure specular reflection (Figure 6 b). A moderate scattering has been simulated when considering the internal cavity surface to be coated with Alanod MIRO UV C [62], which has been tested to have *R*_*c*_ = 0.9 at the LED wavelengths. The scattering data of Alanod at the working wavelength are not known to the authors. When simulating it we considered the data set provided by the supplier, measured at a longer wavelength, where the FWHM of the scattered rays cone angle around the specular reflection direction is about 5°. The cases comparison in Figure 6 g confirmed the little impact of a small scattering come angle to the fluence rate spatial distribution. Also a more diffuse scattering has been simulated. A possible candidate is Polytetrafluoroethylene (PTFE) [63] which is reported to have a reflectivity of about 0.95 at 275 nm and a Lambertian scattering distribution, i.e. all incident rays are diffused with equal probability anywhere in the unit semicircle independently from the incidence angle. The knowledge of the scattering angular distribution is important to properly simulate the filter efficiency and light leak from the apertures.

**Table 1.** Parameters space of the optical simulations to calculate the filter power density distribution. Second last column contains the values of the reference case, last column indicates the range of parameter values variation of the optical simulations

| Object | Parameter | symbol | Unit | Value |  |
| --- | --- | --- | --- | --- | --- |
|  |  |  |  | Reference | Comparison cases |
| Filter | shape |  |  | Cylinder |  |
| | diameter | $D_c$ | mm | 40 | 30, 50, 60 |
| | length | $L_c$ | mm | 100 | 50,150 |
| | reflectivity | $R_c$ | | 0.9 | 0, 0.8, 0.85, 0.95, 0.99 |
| | scattering cone angle | $Sc$ | | 0 | FWHM 5°, Lamb |
| Flange | shape |  |  | Cylinder |  |
| | diameter | $D_f$ | mm | 21 | |
| | length | $L_f$ | mm | 20 | 40 |
| | reflectivity | $R_f$ | | 0 | 0.9 |
| LED | emission power | $I_0$ | mW | 100 | |
|  | Number |  |  | 15 |  |
| | Emission cone angle | $\alpha$ | degrees | 150 | 30 |

The filter flanges are supposed to be composed by an UVC absorbing material. Aside from connecting the filter to the air pipes, the flanges act as reducers of light leak through the holes, blocking all the rays that hit their surfaces. In principle, a narrow angle source and a perfectly plane surface would would not let light exit the filter, for a flange length that can be defined by simple geometrical calculations, as for the blue ray in the sketch of Figure 1. In reality, scatter from internal surfaces will always happen and some rays will leak out. The power density inside the flanges volumes is quite small, if compared to the intensities inside the cavity (Figure 6 j,k). Using a reflective surface for the flanges would increase the overall filter efficiency but at the same time it would increase also the light leaks outside the filter.

## 3. Optical simulations

For this work, the optical ray-trace modeling has been carried out using Zemax OpticStudio^®^, which is a well-known software for optical systems design and analysis. Some works used ray tracing to estimate the performance of UVGI devices such as decontamination chambers [64,65] or to evaluate the fluence distribution for different surface reflectivities and lamp configurations [66]. Analytical formulas to evaluate fluence rate after multiple reflections [44, 67] are not accurate when dealing with omnidirectional light as in the presented work. The goal of the optical simulations is to evaluate the fluence rate distribution inside the filter and the light leak outside the filter holes as a function of some parameters, listed in Table 1. The cavity is designed as a cylindrical pipe with square apertures where the LED packages, a non reflecting square surface of 6 mm side, are inserted as shown in Figure 3. LEDs are arranged in arrays of 5 over a common plate. Three arrays are disposed symmetrically at 120° around the filter axis for a total of 15 sources and 1.5 W emission power. LED positions around and along the optical axis (z axis) ensure symmetry of the fluence rate inside the filter volume. The LED dome, a Quartz spherical window that protects the chip, is designed as a lens with no optical power. In the simulations, the LED dome has then only the sfunction of refracting and reflecting the rays hitting its surface, using the optical parameters from Ref. [68]. For the case of reduced emission angle LEDs (Section 2.2), the micro-lens is simulated as an additional thin spherical Quartz shell posed over the standard device. This solution permits to override the possible lack of information on the micro lens parameters (curvature radii, thickness, conic constants) and still consider the optical effects for the incoming rays. Differently from other works, where the LED intensity variations across the polar angle is defined by mathematical functions [25, 69], ray-tracing permits to easily set the relative radiant intensity, the power emitted from a point source into a small solid angle *8a* about a given direction from the source (with units of *W*/*sr*), and replicate very accurately the vendor datasheet or laboratory measurements.

**Fig. 3.**
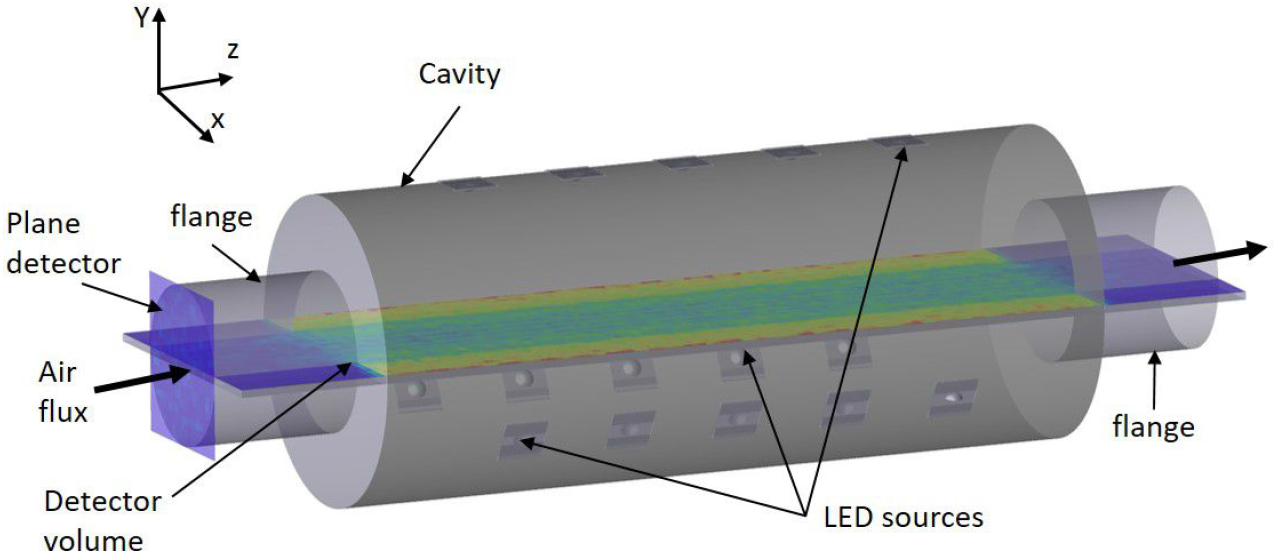
Position of the detectors used to calculate the filter performance in terms of fluence rate (detector volume) and light leak from the cavity to the flanges and outside the filter holes (plane detector). The detector volume is rotated around the z axis to scan the all the volume inside the filter.

### 3.1. Parameters space

The reference parameter values of the filter, listed in Table 1, have been chosen to be reasonable values and are used to compare how the parameter variations impact the fluence rate intensity and spatial distribution inside the filter, and consequently the air sanification efficiency. Cross variations of these parameters have not been simulated. Some parameters have been maintained fixed in the simulation: LED emission power is a multiplicative factor in the fluence rate calculation and can be easily retrieved when varying this parameter; a different number of LED with the same total emission power could slightly impact the power density distribution along the z axis but, for high reflectivities the sources configuration has little impact on the performance [44]; the flange diameter is considered to be 21 mm, compatible with standard breathing circuits; the filter shape is cylindrical, some considerations on different shapes are outlined in Section 2.1.

### 3.2. Observables

The performance of the optical simulations has been evaluated by means of ‘detectors’, which are plane or volume arrays that record the power of all the rays passing through the surface or volume, hence measuring the irradiation of the fluence rate. Their positions are shown in Figure 3. The plane array is at the end of one of the two flanges with its surface perpendicular to the filter axis. The goal is to measure the amount of radiation exiting the cavity in the different simulated cases. Clearly the same would happen at the other flange but, in the assumption of symmetry, the amount is assumed to be the same. The fluence rate inside the cavity (and the flanges) is measured by a volumetric detector, a three-dimensional array formed by cubic voxels, each one of 1*mm*^3^ in order to properly sample the fluence rate spatial variations. The detector volume dimensions are defined by the filter length, the filter maximum diameter while the third dimension is 1mm. The measurement of the volume power density for the whole internal filter volume is achieved by rotating the detector volume around the z axis and recording data for a certain number of angles. In this way it is possible to calculate the mean fluence rate, assumed to be symmetric around the optical axis, for different radial distances, as displayed in the images of Section 3.4.

### 3.3. Volumetric power density

Ray-tracing programs compute the rays path by applying the Snell’s law at the interface between different mediums and the Beer-Lambert’s law for the energy absorption, when applicable. For any source, the emission power and emission angle distribution are split in a finite number of emitted rays in different directions. Every ray carries the energy amount given by the source total energy divided by the number of rays. Thus, a simulated ray is assigned the energy corresponding to a certain number of photons or to a certain number of moles (6.0221418 × 10^23^) of photons [56].

Fluence rate definition of Section 2 applies to infinitesimal volumes. For finite volumetric cells the proposed approach is to consider that the ray power is artificially confined into a 1D polygonal path axis and can be described by a linear power density. The contribution of each ray to the volumetric power density is calculated by integrating the linear power density over the path *l* within the considered cell and dividing by the cell volume *V*_*eell*_. Figure 4 a shows three rays crossing a cubic cell. The rays paths inside the volume are the red segments and can have different lengths. Assuming *p* being a linear ray power when entering the cell, the linear power *p*_*l*_ for the path *l* inside the cell can be written as:

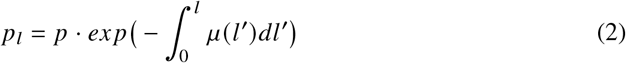

where *µ* is the attenuation coefficient of the medium. In this paper *µ* is set to 0 since the UV air absorbance can be considered negligible [70, 71]. Also the possible light refraction by the aerosol particles, which would vary the rays path, has not been considered for simplicity, since its effect is quite small when considering air respect to surfaces [72]. The single ray contribution to the volumetric power density *p*_*V*_ is:

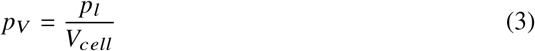

**Fig. 4.**
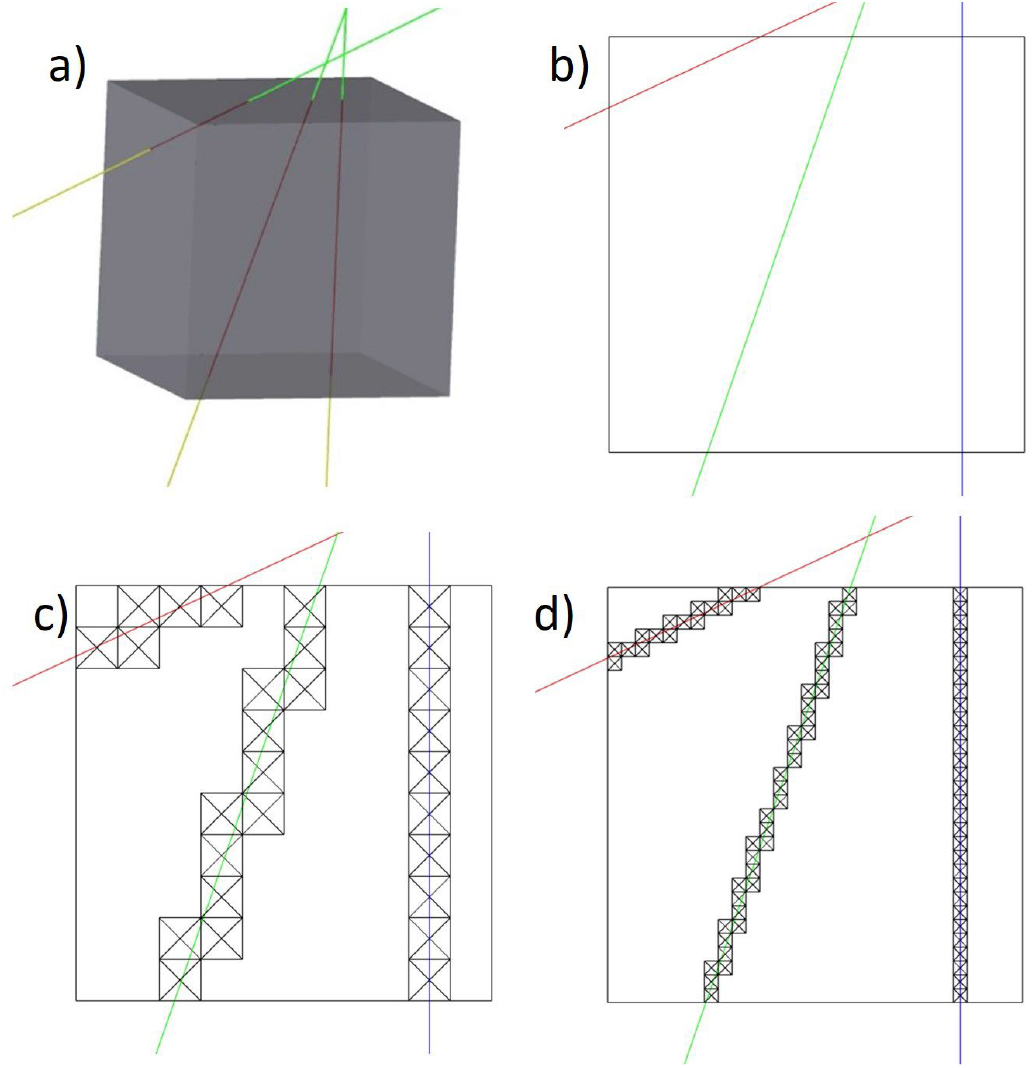
a) Cubic volume passed by three rays. The path of each ray inside the volume is defined by the respective red segment and depends on the ray’s incidence angle and position respect to one of the cube faces; b), c), d) projections of the cube which is crossed by three rays with different arriving angles and different paths inside the volume. The three images represent how the sampling of the cube with a different number of volume elements affects the ray’s path determination and consequently the calculation of the volume power density.

The total volumetric power density *P*_*V*_ is the sum of all *p*_*V*_.

#### 3.3.1. Method for fluence rate estimation

In Zemax OpticStudio, the volumetric power density is calculated numerically by using the “detector volume” object, a parallelepiped volume where size and number of voxels, smaller parallelepiped volumes, are defined by the user. The power of each ray entering one of the faces of each voxel is recorded, without considering the actual path inside the considered volume. By defining each voxel as a cube, each ray path inside the voxel can be considered as long as the cube side, no matter the entering direction. Each voxel total power is then multiplied by the side length and divided by the detector volume. The total volume power density is the sum of all voxels contribution. In the simplified case where all the rays are parallel to each other, i.e. the light is collimated, and enter the volume perpendicularly to a face, the ray’s power multiplication to the voxel side is absolutely correct. The path of each ray has exactly the length of the voxel side. Each layer inside the volume, parallel to the entrance face, is crossed by the same flux, if absorption is not considered as described in Section 3.3. Statistically, also for the opposite case respect to the collimated light, which is the isotropic rays direction distribution as for a Lambertian scattering of the internal reflective surfaces of the cavity, the rays mean path inside the cube volume can be approximated to the side length [73]. For a sufficiently high number of rays, the volume power density can be calculated as the total linear power entering each cube face multiplied by the cube side length and divided by the volume. In the other cases, the calculated power density by ray-tracing could be affected by a discretization problem, because the ray contributions to the total power are not weighted by their path inside the measuring volume. Figure 4 b shows the projection of a detector cube crossed by three rays. Blue ray is perpendicular to the cube face, red and green rays are skew. Assuming that all the rays have the same power, they contribute equally to the volume power density calculation in the ray-tracing program, even though their paths inside the volume are different as their actual linear power density contribution as indicated in Section 3.3. By increasing the sampling, as shown in Figure 4 c, d, it is possible to mitigate the voxel discretization and approximate the calculated path to the real rays paths.

The unit of measure of the volumetric power density *P*_*V*_ is *W* / *m*^3^, while fluence rate (*W*/*m*^2^), is usually used when referring to air sanification doses. The authors agree with Kowalski, who in chapter 3 of the ‘Ultraviolet Germicidal Irradiation Handbook’ [15], stats that “irradiation field and microbial mass absorbing the media exist in a volume and irradiance should have units of *W*/*m*^3^”. Moreover, the air flux is measured in litres (or *m*^3^) over time and the sanification rate is given in terms of CFU (Colony Forming Units) reduction over *m*^3^. Anyway, to be congruent with the pathogen inactivation doses units of measures, *W*/*m*^2^ *time*, the volumetric power density values from the optical simulations are transformed in surface power density *P*_*A*_ from rays entering the cube through each of the six surfaces with area *A*, using the following relationship:

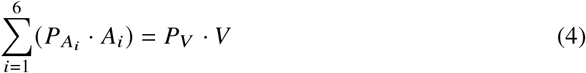

We checked that the total power of six plane detectors placed over the faces of a cube has the same value as the volumetric power of one detector volume with the size of the cube, only the unit of measure are different. The values are exactly the same in the case of collimated of isotropic light distributions, and in the other cases the values tends to converge with a good approximation when increasing the sampling. Fluence rate *F*_*r*_ is the sum of the six surface power densities 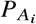.

### 3.4. Trade-off analysis

This section presents the results of the optical simulations regarding the filter performance, for different parameters variations listed in Table 1. The intensities in each image of Figure 6 in this section are normalized, in order to better understand the dependence to a particular parameter. It can be noticed that, even though the sources emission power and number are the same for every case, the total fluence rate inside the filter volume is different also in cases with the same geometrical size. This is due to the mean rays path length inside the cavity, which can be different. After each reflection a ray loses a fraction of its energy, thus the longer the (mean) path between two different surfaces hit, the bigger the contribution to the overall fluence rate. In the following the parameters variation effect on the fluence rate intensity and spatial distribution are discussed.

### Cavity reflectivity

The *R*_*c*_ parameter has a fundamental importance in the filter sanification efficiency. The curve in Figure 5 shows an exponential increase of the fluence rate with *R*_*c*_. Anyway, light leaks from the cavity, to the non reflective flanges or the non reflective LED packages or through the filter apertures, decrease the theoretical exponential growth described in Section 2.3. However, the reference *R*_*c*_ = 0.9 produces a fluence rate enhancing factor of 5 times respect to the case of non reflective cavity surface. Smaller cavity diameters are subject to a relative slower growth due to the higher light leak values and the higher percentage of non reflecting area inside the cavity.

**Fig. 5.**
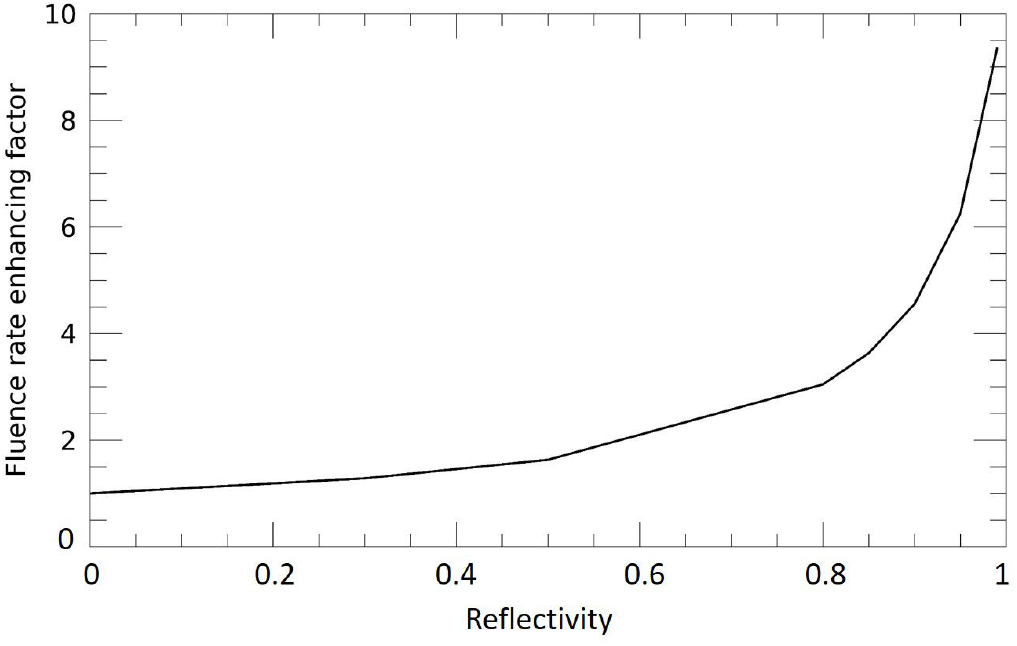
Fluence rate enhancement as a function of the cavity internal reflectivity. The values are normalized to *R*_*e*_ = 0, where the fluence rate emitted by the source, before being absorbed by the filter surfaces, is considered.

### Cavity diameter

As described in Section 2.2 and shown in Figure 2 d, the *D*_*c*_ parameter varies the radial fluence rate distribution and the optimal diameter should be chosen considering both the source emission angle and the air flow distribution, as explained in Section 5.

### Cavity length

The parameter *L*_*c*_ produces a reduction of the mean fluence rate close to the filter axis since there is less rays overlap from the sources, which are positioned more distant one anothe as the cavity length increases. Sources positioning is of little important in the presence of highly reflective surfaces [44], since it can modify the axial fluence rate intensity profile but the integrated value along any axial path is not varied in an appreciable way. For the considered flux value, where air velocity is constant inside the filter, the air residence time is proportional to the filter length and thus the smaller fluence rate mean values for longer cavities are counterbalanced by the increased air residence time. The fluence slightly increases as the cavity length increases.

### Source emission angle

Figure 6 i demonstrates how the sources emission angle determines the fluence rate intensity as a function of the radial distance, as explained in Section 2.2 and displayed in Figure 6 d. For the wide angle emission of the reference source, the power density is a radial increasing function, while for the narrower angle emission source the fluence rate decreases with the radial distance. For a given cavity diameter, there is an emission angle pattern that maximises the fluence rate in function of the radial distance, to be combined to the air flux velocity field.

**Fig. 6.**
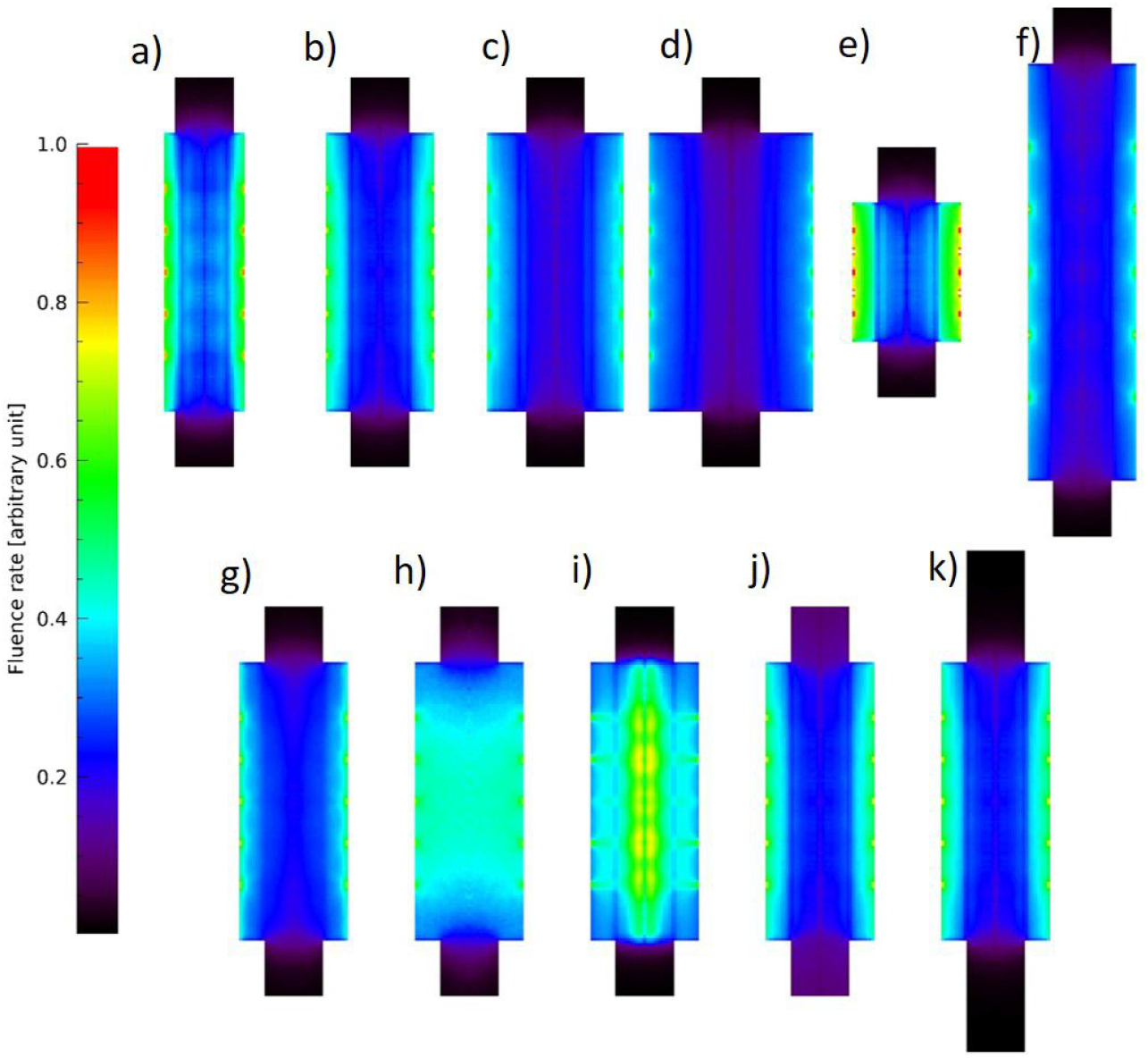
fluence rate inside the filter for different parameter variation. Intensities are normalized. The reference case is figure b. The other displayed cases are: cavity diameter variation in figures a,c,d; cavity length variation in figures e,f; Scattering variation in figures g,h; source emission cone angle in figure i; flange reflectivity in figure j; flange length in figure k.

### Scattering

As shown in Figure 6 b, g, the cases with no scattering and Alanod-like scattering give pretty similar results, meaning that when the rays diffusion cone angle after the reflection is relatively small, the fluence rate smoothing is quite limited. The case of pure Lambertian scattering (Figure 6 h) has instead the effect of homogenizing the fluence rate inside the cavity and nullifying any spatial diversity as a consequence of the source emission angle pattern. It applies also to the narrow angle LEDs. Scattering causes also an increase of the light leak.

### Flange reflectivity and length

The flange is considered absorbing to reduce the light leak. The coating of the flange with a reflective surface, *R* _*f*_ = 0.9, increases the fluence rate inside its volume and also the light leak. On the contrary, a longer absorbing flange reduces the leak. The images in Figure 6 j,k show the two simulated cases.

### 3.5. Light leak considerations

UV light has known effects on direct or indirect exposure. For skin one of the most acute effects is the induction of a cascade of mediators in the skin that together causes “sunburn”. UV radiation is also classified as a “complete carcinogen” because it is both a mutagen and a nonspecific damaging agent and has properties of both a tumor initiator and a tumor promoter [74]. Moreover, if the eye is exposed to excessive UV radiation, several severe consequences are likely to take place, including photokeratitis, erythema of the eyelid, cataracts, solar retinopathy, and retinal damage [75]. The threshold limit value (TLV) for human UV exposure is an “effective” (defined on the maximum sensitivity of the human eye, which was found to be at approximately 270 nm [76]) UV dose of 3 *mJ*/*cm*^2^ in an 8 h time frame based on regulation provided by different associations such as European Agency for Safety and Health at Work [77].

The light leak from the filter apertures for the different considered cases are listed in Table 2. Theoretically, if the absorbing flange is long enough to avoid to see, from outside the filter, the cavity reflective surfaces, no light would leak out even in presence of scattering. The case of longer reflective flange demonstrate it. The case with narrow emission angle sources and no scattering has the lowest light leak value since only a very small fraction of rays are emitted at high angles (Figure 2 bottom). Lambertian scattering or the reflective flange cases produce the highest light leak values. The pipes transporting the air to or from the patient, to which the filter should be connected, are usually made of PVC or other plastic material which blocks the UVC light. The pipes can be thought of as very long flanges. A possible concern would rise if, for a specific application, one of the filter apertures irradiated to open air and human exposure were possible. In that case a curved pipe would definitely resolve the issue. It would make the system loose its geometrical linearity and some air load loss could happen but, these considerations, as well as the ageing of the flange or air pipe, will be taken into account for specific application cases.

**Table 2.** Delivered fluence and light leak for the different simulated cases

| Case | Parameter variation<br>respect to reference case | | | Equivalent fluence<br>for $v_{in} = 5m/s$ | Light leak<br>through each aperture |
| --- | --- | --- | --- | --- | --- |
| | symbol | unit | value | unit<br>$mJ/cm^2$ | Unit<br>$mW/cm^2$ |
| Reference case |  |  |  | 5.46 | 5.25 |
| 1 | $R_c$ | | 0 | 1.42 | 0.15 |
| 2 | $R_c$ | | 0.8 | 4.03 | 4.66 |
| 3 | $R_c$ | | 0.85 | 4.63 | 4.93 |
| 4 | $R_c$ | | 0.95 | 6.72 | 5.67 |
| 5 | $R_c$ | | 0.99 | 8.44 | 6.11 |
| 6 | $D_c$ | mm | 30 | 7.29 | 6.92 |
| 7 | $D_c$ | mm | 50 | 3.82 | 3.89 |
| 8 | $D_c$ | mm | 60 | 3.03 | 2.96 |
| 9 | $L_c$ | mm | 50 | 2.95 | 5.97 |
| 10 | $L_c$ | mm | 150 | 7.00 | 4.59 |
| 11 | $L_f$ | mm | 40 | 5.46 | 1.47 |
| 12 | $R_f$ | | 0.9 | 5.89 | 31.01 |
| 13 | $Sc$ | mm | 5 | 5.42 | 6.79 |
| 14 | $Sc$ | | Lambertian | 8.84 | 11.64 |
| 15 | $\alpha$ | degrees | 30 | 10.77 | 2.32 |

Symbols legend: $R_c$ cavity reflectivity; $D_c$ cavity diameter; $L_c$ cavity length; $L_f$ flange length; $R_f$ flange reflectivity; $Sc$ Scattering distribution; $\alpha$ LED emission cone angle

### 3.6. Possible fluence rate experimental measurement

The experimental measurement of the fluence rate inside the filters is not an easy task since light is expected to be almost omnidirectional. UVC radiometers can measure light arriving over a certain solid angle applying the cosine correction to the recorded data. By placing the radiometer probe in different positions inside the filter volume, entering also through test holes of the cavity,the comparison with the expected intensities in the same positions would be a possible confirmation of the reliability of the optical simulations. Another possible fluence rate measurement can be done by means of chemical actinometry [78–80], such as iodide/iodate enclosed in quartz spheres or tubular [49, 81], but the size of about 10mm limits the resolution, since the presented filter has comparable dimensions. Micro fluorescent silica detector (MFSD) have a much smaller size [82] and could help to better discriminate the fluence variations inside the filter. For these reasons, either a bigger scale prototype is constructed and the fluence rate measurements are done with a better resolution or the verification is done directly on the sanification performance by using a similar set-up as other experiments [44, 49, 57, 58, 83, 84].

## 4. Air flow simulations

The Computational Fluid Dynamic (CFD) analysis has been performed using the Ansys CFX5 software. The elements are all hex elements with dimension of 2 mm and the material is air at 25° C. The boundary condition at the inlet is 5 m/s with a turbulence of 5%, the outlet is an opening at the normal ambient pressure. Regarding the working conditions for the assisted ventilation devices the calculations for air flow spatial distribution inside the filter, to be combined to the fluence rate results, have been done for 100*l*/*min*, corresponding to about *v*_*in*_ = 5*m*/*s* through a tube with the flange diameter. Streamlines of Figure 7e support the hypothesis of laminar flow, where the air trajectories can be considered independent. Also slower fluxes, corresponding to 50 and 20 l/min, have been investigated. The flow is still laminar and substantially there is the same velocity field distribution, the velocities are just scaled down. The velocity field is more influenced by the cavity diameter, since wider cavities reduce more the air velocity and turbulent regions appear close to the cavity walls. Figure 7a-d shows the the velocity field on the longitudinal cross section inside the filter for *v*_*in*_ = 5*m*/*s* for the different considered cavity diameters. The velocity slows down inside the cavity, and a more prominent reduction appears when increasing the distance from optical axis. Closer to the cavity walls, some air re-circulation is present. Re-circulation zones are not considered in the filter efficiency calculations, described in Section 5.2.

**Fig. 7.**
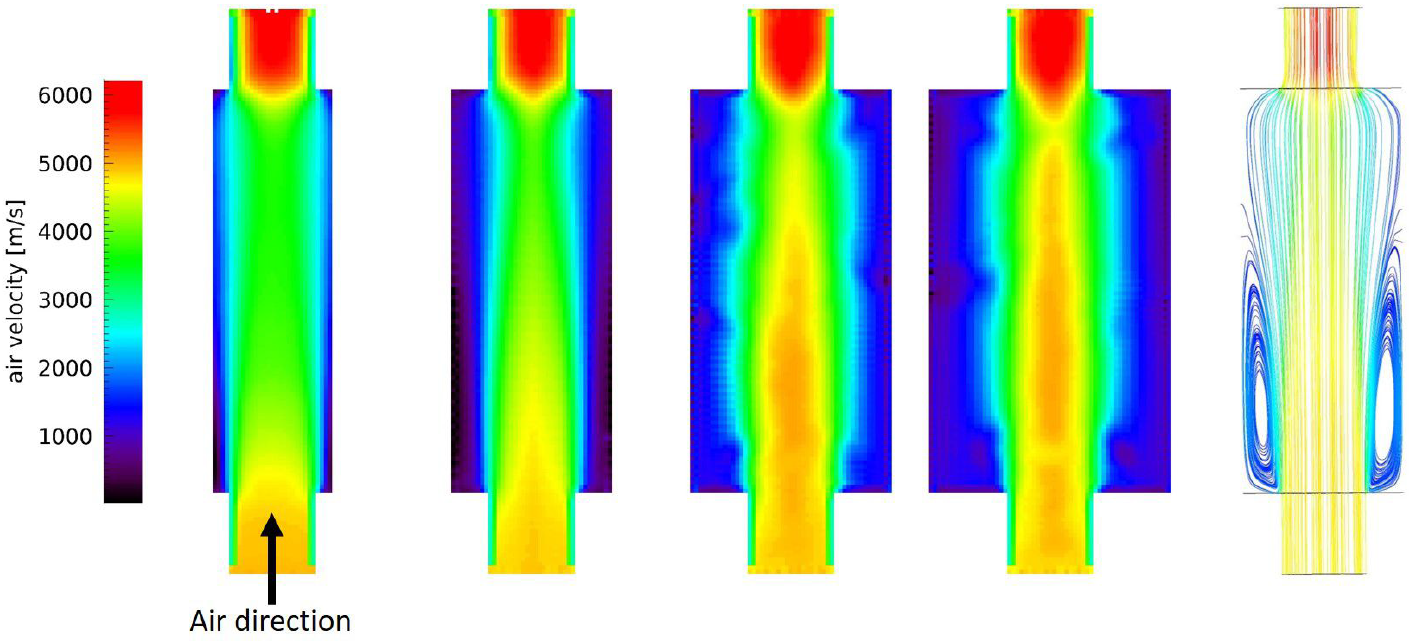
a) to d) Longitudinal section of the filter where the air velocity field is displayed for different cavity diameters, *D*_*c*_ = 3030, 40, 50, 60 mm respectively. Entrance air velocity *v*_*in*_ is 5 m/s and the flow is from the bottom to the top of the images; e) streamlines of air inside the filter for *D*_*c*_ = 40 mm where the laminar flow of air, apart the turbulent lateral regions close to the entrance flange, is evident.

## 5. Filter sanification performance

### 5.1. Inactivation model

Inactivation of the microorganisms is a function of the total UV energy absorbed. A common model for describing microbial inactivation [15] is the exponential relationship:

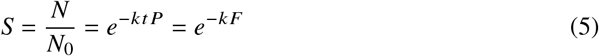

where *S* is the survival fraction of microorganisms after being exposed UVC light, *N*_0_ is the number of survived microorganism before the UVC exposure, *N* is the number of microorganism after the UVC exposure, *k* is the specific rate constant unique to each type of microorganism (*cm*^2^/*mJ*), *P* is the fluence rate, *t* is the time interval and *F* is the fluence (*mJ*/*cm*^2^). On a logarithmic scale, microorganisms decay rate is linear with fluence for values usually starting from *D*_90_, which corresponds to 90% of inactivation rate. The described processes produce the so called shoulder effect in low fluence region of the inactivation curve, where the pathogens decay is almost constant and becomes exponential after a threshold [15]. Below that threshold the survival curve can be described by different models: the two stage inactivation curve, due to the presence of a fraction of microorganisms with a higher level of resistance; the shoulder correction effect in low fluence region, where the pathogens decay is almost constant and becomes exponential only after a threshold [85, 86]. For high enough fluence values, the product high-intensity fluence rate short-time irradiation has the same sterilization effect as the low-intensity fluence rate long-time irradiation [87].

### 5.2. Expected UVGI efficiency

The fluence rate results from Section 3.4 have been combined with the air flow simulations to obtain the fluence and consequently the survival fractions *S*. On the basis of the optical and CFD simulations, the following assumptions and simplifications have been assumed. Air has a laminar flow and there is no air mixing inside the filter, so each path is independent from the others. The regions with air recirculation have not been considered, this assumption is conservative; due to the small fluence rate, the regions inside the flanges are not considered in the dose calculations, except for the high reflective flange case.

Given the filter parameters, the microorganism rate constant *k* and the entrance velocity of air:

- the velocity field from CFD simulations is transformed to a residence time field and multiplied by the fluence rate spatial distribution to obtain the locally delivered fluence in each cell;
- a particle tracking is computed, starting from a grid of positions at the entrance plane of the filter. The air flow is divided in *n* axial paths, discretized as thin pipes with square cross section and composed by *n*_*z*_ small volumes, which follow the particle trajectories;
- for the *n* − *th* path *S*_*n*_, the accumulated fluence across the filter, the amount of UVC energy received by the pathogens in their trajectories inside the filter, and consequently the survival fraction is calculated:

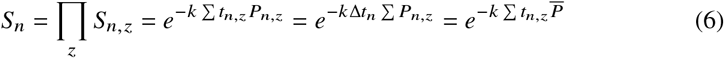

where the subscript *z* refers to the *z* − *th* volumetric element along the path, f..*t* is the mean residence time and 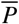 is the fluence rate mean value along the path. In case of non constant velocities, the last or second-last equalities of eq. 6 are not correct but are a good approximation if local velocities of a given trajectory are not very different.
- the survival fraction *S*_*v*_ for a given velocity is the sum of all the paths survival fractions *S*_*n*_, weighted by the pathogen fraction *a*_*n*_ that entered the path.

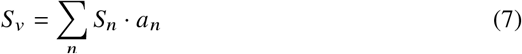

The comparison among the survival rate for the different paths can help to identify the areas of low performance in the filter. These regions can be minimized by changes in the design. For example, in regions with low fluence rate the LEDs can be rearranged to generate a better distribution.

The survival fractions delivered by the filters are calculated by a combination of local fluence as described above. It is useful to introduce the equivalent fluence *F*_*eq*_ as the fluence that delivers the same survival fraction *S*_*v*_ as the filter at a given air entering velocity, calculated using the above steps.

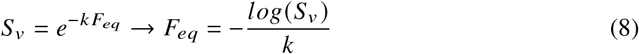

Table 2 presents the *F*_*eq*_ for the different filter configurations.

There is often some discrepancy among the inactivation doses for the same pathogen since methods and test set-ups are not standardized. Some works list the constant rates *k* [85] but more often the measurements report the doses for one or more logarithmic inactivation rates [23] and the “k” constant must be retrieved. Regarding the case of SARS-Cov 2, a review on coronavirus family inactivation doses and experimental data comparisons on SARS-Cov 2 [42, 88, 89] agree that about 3-4 *mJ*/*cm*^2^ are sufficient to produce at least a log3 inactivation rate. The expected fluence values of the different simulated filter cases are listed in Table 2. Apart from the cases with non reflective surfaces (case 1) and short cavity length (case 9) the equivalent fluences are always larger than the target values. Note that the SARS-Cov 2 doses are given for plate measurements. Air sanification could be more effective since light arrives from all directions and also the shading or shielding effects that produce the tail on the lower end of the typical survival fraction curve, which limits higher log inactivation, could disappear [16, 72]. Results from Table 2 point out the directions for the most effective filters, respect to the reference case: high reflectivity, small cavity diameter, optimized source emission angle and longer cavity length. The delivered fluence when combining some of these parameters could increase significantly and also more resistant microorganisms as bacteria or spores [15] could undergo relevant inactivation rates. The case with high reflective flange produces a small delivered fluence increase at the price of a significant increase of light leak. The Lambertian scattering of the cavity surface increases the filter efficiency respect to the reference case but reduces it for the small emission cone angle case since fluence will be almost uniformly distributed inside the cavity volume. Bigger cavity diameters are less effective but in the simulations the turbulent fraction of the air flow has not been considered, thus their delivered fluences are under estimated.

## 6. Conclusions

This paper investigated the performance of a compact filter for air sanification using UVC light from commercial LED sources, with the focus on concrete applications, as the exhaled air from assisted ventilated hospitalized patients. This filter results to be highly effective to inactivate SARS-CoV 2, also considering the maximum air flux values for the assisted ventilation devices. Lower fluxes or optimized parameters combination would permit to apply this concept to more resistant pathogens. The innovation is the quantitative and accurate analysis of the performance of a device that exploits the concept of enhancing fluence rate produced by multiple light reflections inside an optical cavity. Ray-racing and CFD analyses are fundamental to have a realistic and accurate estimation of the performance, since analytical formulas to estimate source emission power and spatial irradiation could not be precise enough since in the internal volume of the filter the light is emitted, reflected and scattered in all directions. The fluence rate calculations have been carried out by considering volumetric units and successively transformed into surface units to be consistent to the standard units of measures. Simulation results show that there is a dependency on some filter parameters which control the spatial distribution and intensity of the fluence rate. There are many combinations of the filter parameters that have not been studied since the goal of the optical simulations trade-off was to understand the impact on the performance when varying a single parameter respect to the reference case. Anyway, the way to achieve the best performance is clear: use materials with the highest possible reflectivity, sources with an emission cone angle tailored to the cavity diameter and arrange the sources in such a way to increase irradiation of the portions of the cavity where air flow is faster.In the air stream velocity conditions considered in this paper, the wide angle emission LEDs appear less effective respect to narrower cone angle sources. By inserting a deflector at the entrance of the cavity to to force air to pass close to the walls, where these sources produce the maximum fluence rate, would highly increase the performance, shifting the mechanical complexity from the source to the cavity. Moreover, the filter shape can be studied to increase the efficiency, by increasing irradiance where air is faster, and reduce light leak. And clearly, when more powerful and more efficient UVC LEDs will be available, the delivered fluence will increase proportionally with the power, and another step will be done toward the increase of the pathogen sanification effectiveness. The same analysis described in this paper can be applied to air sanification in industrial or domestic air ducts, which is another relevant issue raised during the current pandemic. The much bigger air volumes to be processed require, at the moment, the use of mercury lamps but a proper system design would reduce the number of sources with the same sanification efficiency. In conclusion, some further work is still necessary to optimize the filter design but already now it can be said that the LED era, at least as far as air sanification effectiveness is concerned, has already begun.

## Data Availability

no data have been used

## Acknowledgments

The authors gratefully acknowledge Prof. Claudio Borghi for and Dr. Andrea Zanoni for their support and for providing help on the link between the technical work and the medical applications.

## Disclosures

The authors declare that they have no competing interests.

## Notes

### Competing Interest Statement

The authors have declared no competing interest.

### Funding Statement

no external funding was received

### Author Declarations

no ethics committee was necessary for this paper

